# Therapeutic Alliance Predicts Clinical Outcomes in a Fully Virtual Telepsychiatry Practice

**DOI:** 10.1101/2025.05.09.25327264

**Authors:** Nathaniel D. Phillips, Georgia Gaveras, Cheryl Person

**Affiliations:** Talkiatry, New York, NY, USA

**Keywords:** Telepsychiatry, PHQ-8, GAD-7, telehealth, depression, anxiety, therapeutic alliance, WAI-SR

## Abstract

**Background:** Given the increasing demand for accessible mental health services, fully virtual telepsychiatry has become a vital component of modern health care delivery. Therapeutic alliance, the collaborative and affective bond between patients and therapists, is a well-established predictor of clinical outcomes in traditional face-to-face psychotherapy. However, the relationship between therapeutic alliance and clinical outcomes in an outpatient telepsychiatry setting remains less understood.

**Objectives:** Our primary objective was to evaluate the relationship between therapeutic alliance and clinical outcomes for depression and anxiety in an outpatient telepsychiatry practice.

**Methods:** This retrospective cohort study analyzed data from 201 treatment seeking adults receiving outpatient telepsychiatry care. Treatments included a comprehensive psychiatric evaluation, supportive psychotherapy and medication management conducted by a psychiatrist utilizing a fully virtual platform. Anxiety and depression symptoms were assessed using the Generalized Anxiety Disorder scale (GAD-7) and Patient Health Questionnaire depression scale (PHQ-8). Eligible participants completed a baseline clinical assessment within 1 week of their first visit that revealed at least moderate baseline symptoms (PHQ-8 and/or GAD-7 ≥10). Therapeutic alliance ratings from the patients’ perspective were collected using the Working Alliance Inventory-Short Revised (WAI-SR). Follow-up clinical assessments were conducted between 8 and 16 weeks after their first visit. We used logistic regression models to assess whether therapeutic alliance was associated with clinically significant improvement, which was defined as having ≥50% improvement in depression and/or anxiety symptoms at follow-up after adjusting for demographics (age, region, sex, insurance type, urban/rural) and baseline clinical scores (PHQ-8 or GAD-7).

**Results:** Among 201 patients, mean baseline symptom severity scores for depression and anxiety were 14.90 (SD 3.81) and 14.64 (SD 3.39), respectively. In response to telepsychiatry treatment, patients showed significant reductions in anxiety symptoms with a 41.1% improvement in symptoms from baseline (*d*=1.16, p<0.001) and a 38.8% improvement in depressive symptoms from baseline (*d*=1.07, p<0.001). Importantly, after controlling for other covariates, higher therapeutic alliance scores were significantly associated with greater likelihood of clinically significant improvement for both anxiety (OR=1.08, p<0.001, 95% CI: [1.04, 1.14]) and depression (OR=1.05, p=0.01, 95% CI: [1.01, 1.09]).

**Conclusion:** Therapeutic alliance scores independently predicted meaningful clinical improvements in an outpatient telepsychiatry setting. These findings speak to the importance of fostering strong therapeutic relationships between patients and psychiatrists, even when treatment is delivered virtually.

## Introduction

Depression and anxiety are among the most common mental health disorders globally, collectively contributing to significant personal, social, and economic burdens^1–3^. In the United States alone, the economic burden of depression exceeds $300 billion annually, surpassing many other chronic medical conditions such as coronary heart disease^4^. Despite the prevalence and economic impacts of these disorders, many individuals face barriers in accessing timely and effective mental health care due to geographic disparities, provider shortages, and insurance limitations^5–7^.

Telepsychiatry and teletherapy have emerged as promising alternatives to traditional face-to-face mental health care, addressing many of the aforementioned barriers by providing broader access, cost-effective treatment, and convenience for patients^8–14^. Prior studies have demonstrated that fully virtual psychiatry care leads to significant and clinically meaningful reductions in depressive and anxiety symptoms, comparable to outcomes achieved with in-person treatment ^10,15^. Moreover, mental health professionals have expressed positive attitudes toward telepsychiatry, recognizing its potential as an effective treatment delivery model beyond pandemic-specific contexts ^16,17^.

A crucial component contributing to positive treatment outcomes in both traditional and virtual mental health care is therapeutic alliance, which is defined as the collaborative, goal-oriented bond established between the patient and therapist ^18–21^. Therapeutic alliance, especially from the perspective of patients, is one of the most robust predictors of clinical outcomes for both face-to-face and fully virtual therapy^18,22^. Furthermore, therapeutic alliances formed between patients and therapists in teletherapy settings can be equivalent or better than those established during in-person therapy, highlighting the utility of virtual platforms in facilitating robust therapeutic relationships^19,23,24,25,26^.

Despite the demonstrated efficacy of telepsychiatry and the established significance of therapeutic alliance in therapy settings, studies of the impact of therapeutic alliance on clinical outcomes within real-world telepsychiatry practices have been limited^27^. To our knowledge, there is no existing research demonstrating the impact of patient-reported therapeutic alliance on clinical outcomes in a fully virtual telepsychiatry practice. Addressing this gap is essential for guiding clinical best practices and advancing evidence-based care, particularly as fully virtual psychiatric treatment emerges as a cornerstone of mental health service delivery.

### Objectives

The primary objective of this study was to evaluate the relationship between therapeutic alliance from the patient perspective and clinical outcomes among patients receiving psychiatric care through an outpatient telepsychiatry practice. We hypothesized that a stronger therapeutic alliance, measured after the patients’ second visit, would predict clinically meaningful improvements in symptoms of depression and anxiety.

## Methods

### Study cohort and treatment strategy

The study population was comprised of adults seeking treatment through an outpatient telepsychiatry service. Patients initiated care by registering and completing a short screening questionnaire at publicly available online scheduling platforms or the service website. Following screening, patients selected a psychiatrist after reviewing profiles that contained psychiatrists’ photos, clinical specialties, and appointment availability. Patients then scheduled an initial evaluation session with their chosen psychiatrist. All patients were asked to complete baseline assessments of depressive and anxiety symptoms within one week prior to their first visits.

Depressive and anxiety symptoms were assessed using the Patient Health Questionnaire-8 (PHQ-8) questionnaire and the 7-item Generalized Anxiety Disorder (GAD-7) questionnaire, respectively^28,29^. Subsequent clinical assessments were conducted at monthly intervals.

Therapeutic alliance between patients and their treating psychiatrist was measured after the second visit using the patient reported Working Alliance Inventory-Short Revised (WAI-SR) ^30^. The final study sample was comprised of patients who: ***(1)*** were at least 18 years of age; ***(2)*** completed baseline GAD-7 and/or PHQ-8 surveys within 1 week of their first visit, with scores of ≥10 (moderate baseline symptoms); ***(3)*** had completed a WAI-SR questionnaire after the second visit; and ***(4)*** had completed at least one repeat GAD-7 and PHQ-8 measure 8-16 weeks after their first visit. The timeframe for this study was May 3, 2024 through May 2, 2025.

Patients who completed an initial visit between May 31, 2024, and January 9, 2025, with at least one subsequent visit completed at least 8 weeks after an initial visit, but no later than 16 weeks were eligible for the study. The last possible date for completing follow-up clinical scales and the WAI-SR was May 2, 2025. If a patient completed multiple GAD-7 or PHQ-8 measures 8-16 weeks after their first visit, only the first completed measure was selected for this analysis.

Clinical care, including psychotherapy and medication management, was delivered virtually using a commercially available audiovisual telehealth system. Psychiatrists conducted comprehensive initial psychiatric evaluations and collaboratively developed individualized treatment plans with their patients. Subsequent appointments involved supportive psychotherapy and ongoing medication management, with psychiatrists typically dedicating 20-30 minutes per session to therapeutic interaction. Each patient included in the analysis had at least two psychiatric sessions during the study period. The mean number of visits across patients was 6.75.

### Ethical Considerations

All data in this study were routinely collected as part of standard clinical practice. This study was reviewed and granted exempt status by the institutional review board of Advarra (Pro00084942). A full waiver of Health Insurance Portability and Accountability Act (HIPAA) authorization was granted under an expedited review process. All data were de-identified prior to analyses.

### Measures

Patient outcomes were measured using standardized, self-administered measures of depression and anxiety symptoms. Depressive symptoms were assessed with the PHQ-8^29^, an 8-item instrument widely used in healthcare contexts due to its reliability and sensitivity to clinical change over time. For each item, patients rated how often they experienced symptoms in the prior 2 weeks on a scale from 0 to 3, with 0 indicating “Not at all”, 1 indicating “Several Days”, 2 indicating “More than half the days”, and 3 indicating “Nearly every day”. Total scores are calculated as the sum across all eight items. Total scores classified depression into standard severity categories: minimal (0–4), mild (5–9), moderate (10–14), moderately severe (15–19), and severe (20–24). Anxiety symptoms were measured using the GAD-7^28^, a 7-item questionnaire similarly validated for tracking generalized anxiety disorder symptoms in clinical practice. Like the PHQ-8, each item in the GAD-7 asks patients to rate how often they have been bothered by symptoms in the past 2 weeks on a scale from 0 to 3, with a final score calculated as the sum across all items^1^. GAD-7 scores also categorized anxiety severity into minimal (0–4), mild (5–9), moderate (10–14), and severe (15–21). Patients completed the PHQ-8 and GAD-7 assessments within 7 days prior to their initial psychiatric visit, and every subsequent month during treatment.

Therapeutic alliance between patients and their treating psychiatrist from the patient perspective was measured using the WAI-SR ^30^. This instrument consists of 12 items that evaluate the patients’ rating of the therapeutic alliance with their psychiatrist based on the goals (agreement on the goals of therapy), tasks (agreement on the tasks of therapy) and bond (perception of an affective bond with their therapist). Minor adaptations to the instrument included replacing the word ‘therapist’ with ‘psychiatrist’ and replacing the word ‘therapy’ with ‘treatment’ throughout to more accurately assess the care being provided (see supplementary materials). Patients rated each item on a Likert scale from 1 (Seldom) to 5 (Always). The total score, which is calculated as the sum of all items, ranged from 12 to 60, with higher values indicating higher perceived alliance. The WAI-SR has demonstrated high reliability and validity in both outpatient and inpatient settings^31^. In this sample, the Cronbach alpha for the total score was 0.95.

Demographic variables collected for analysis included patient age group (18-24-years old, 25-39-years old, 40-54-years old, 55-64-years old, or 65+ years old), sex (female or male), US Region (Midwest, Northeast, South or West), urban/rural classification based on patient zip code using USDA classifications^32^, and insurance type (commercial or Medicare).

### Statistical Analysis

Changes in GAD-7 and PHQ-8 scores were calculated by subtracting the follow-up from baseline scores for each patient. Changes in symptoms for GAD-7 and PHQ-8 were conducted in separate analyses and only including those patients with at least moderate symptoms for that symptom at baseline. We used Wilcoxon signed-rank tests to examine whether statistically significant changes in GAD-7 and PHQ-8 were observed among patients. We calculated clinically significant symptom improvement as a binary variable indicating whether a patient had a follow-up score at least 50% lower than that at baseline. We further characterized patients by their degree of improvement using two criteria: ***(1)*** none or minimal symptoms, defined as having a follow-up score less than 10; and ***(2)*** in remission, defined as having a follow-up score <5.

For our primary outcome analysis, we regressed the aforementioned binary clinically significant improvement variables on total alliance within two logistic regression analyses, one for GAD-7 and one for PHQ-8. The analysis included five demographic covariates and one baseline clinical covariate. Due to sample size, no interaction terms were included in the models and patients with missing data were excluded from analyses. Statistical analyses were performed in R ^33^ using tidyverse packages ^34^. Statistical summaries were created using the broom package^35^. Figures were created with the ggplot2 package^36^. Tables were created with the gt^37^ and gtsummary ^38^ packages. Effect sizes for paired samples were calculated with the lsr package^39^.

## Results

### Overview

During the study timeframe, 497 patients completed the WAI-SR and baseline GAD-7 and/or PHQ-8 scales. Of those, 352 had baseline clinical scores of ≥10 in one or both scales indicating moderate or greater symptom severity. Of these, 151 participants did not complete a repeat outcome assessment within the 8-16-week follow-up window, while 201 had at least one repeat clinical outcome measure within the 8-16-week follow-up window. The demographic and baseline characteristics of the patients are presented in Table 1. Compared to patients with only baseline measures, patients with repeat measures were statistically more likely to be older and have Medicare insurance.

**Table 1:**
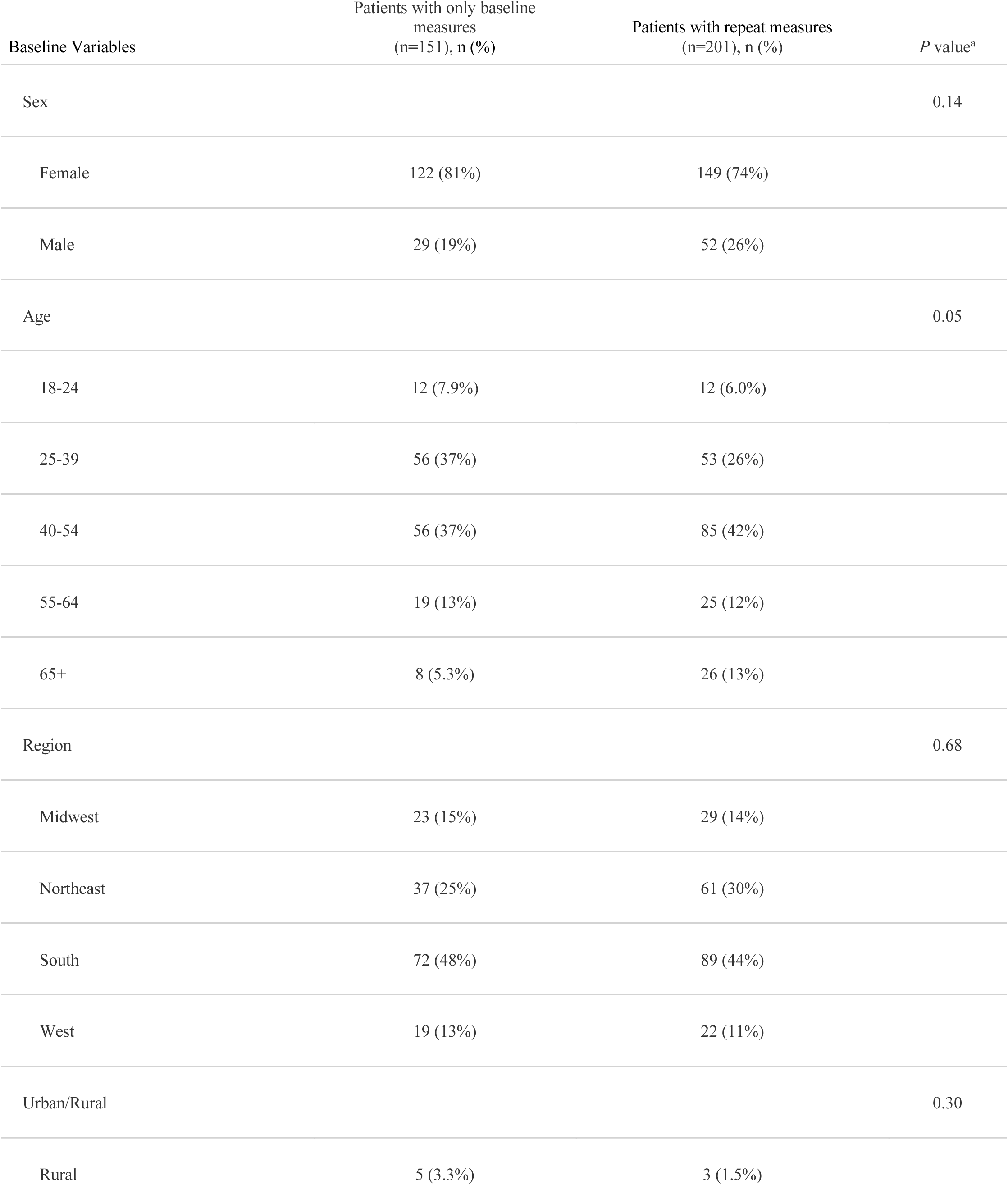
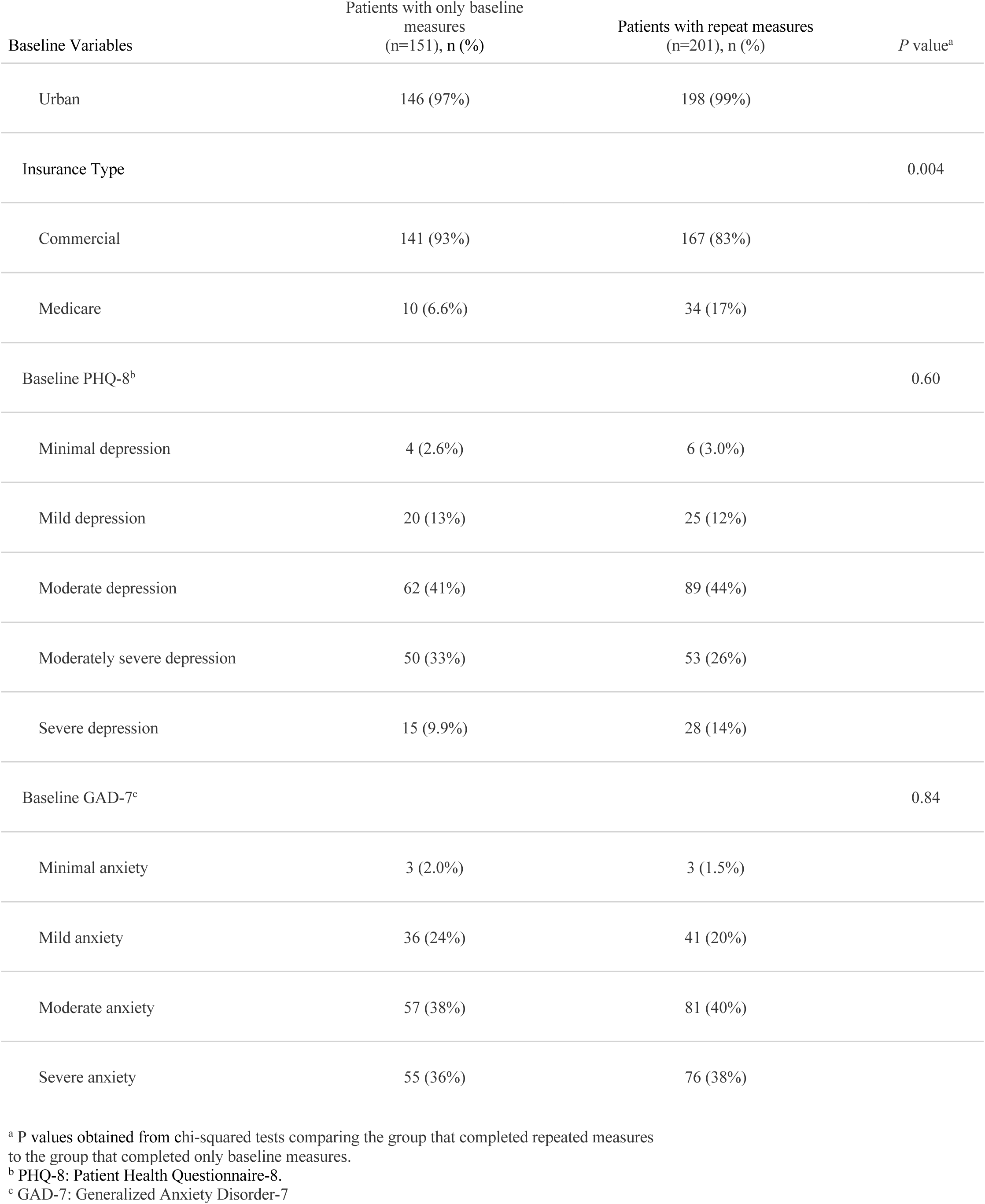
Demographic and baseline characteristics.

### Clinical Change Summary Statistics

Descriptive statistics of clinical status at baseline, follow-up, and changes from baseline to follow-up are shown in Table 2. Patients had mean baseline PHQ-8 and GAD-7 scores of 14.90 and 14.64, respectively, and mean follow-up scores of 9.12 and 8.62, respectively. On average, patients improved their PHQ-8 scores by 5.78 points (38.8% from baseline) and GAD-7 scores by 6.02 points (41.1% from baseline), over the course of 8-16 weeks of treatment. The change from baseline to follow-up was significant for both PHQ-8 (V=12,229.50, p<0.001) and GAD-7 (V=11,271.00, p<0.001). The effect sizes of these changes using paired Cohen’s *d* were large: *d=*1.07 for PHQ-8 and *d*=1.16 for GAD-7. By classifying the degree of improvement for each patient from baseline to follow-up, we found that most previously symptomatic patients improved to having minimal or mild symptoms, (54.7% improvement for PHQ-8 and 62.4% improvement for GAD-7). Furthermore, over 45% had at least a 50% reduction in symptoms (47.1% for PHQ-8 and 46.5% for GAD-7), and over 23% achieved full remission (27.6% for PHQ-8 and 23.6% for GAD-7).

**Table 2:**
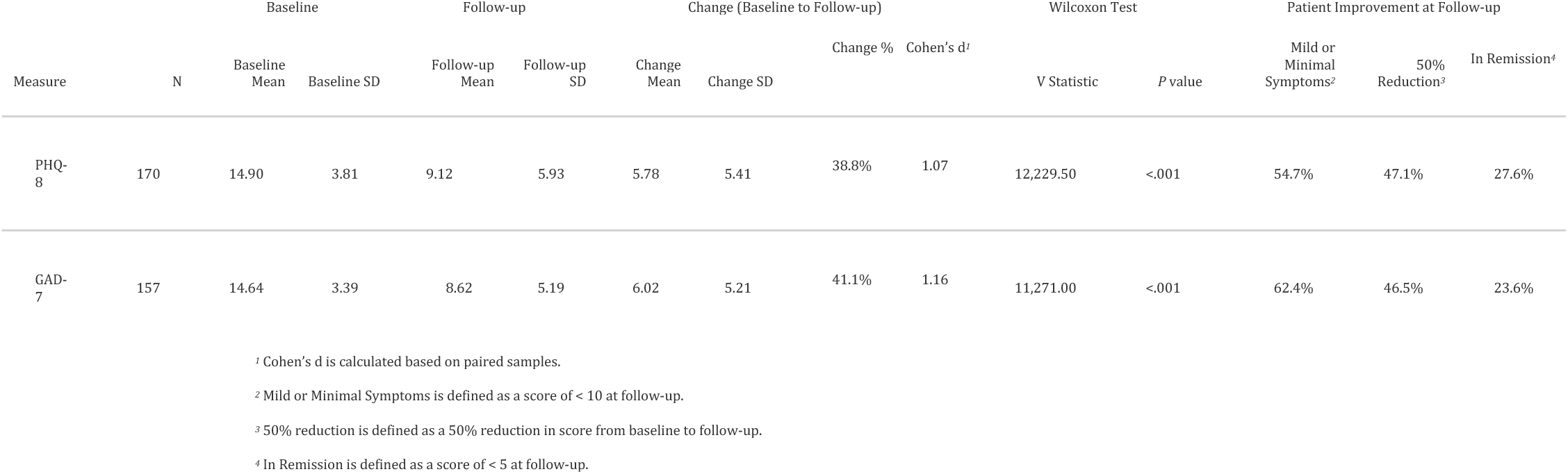
Changes in depression (PHQ-8) and anxiety (GAD-7) symptoms from baseline to follow-up.

### Relationship Between Alliance and Clinical Outcomes

Next, we analyzed the relationship between WAI-SR and clinical outcomes. First, we compared alliance between patients who improve to having mild or moderate symptoms at follow-up to those that did not. Results of the multivariate logistic regression analysis predicted the likelihood that patients improve to having minimal or mild symptoms at follow-up as a function of alliance after controlling for demographics and baseline clinical symptom severity.

Overall, patients indicated high therapeutic alliance with their psychiatrists. The mean WAI-SR was 51.50 out of 60 (median=55.0, SD=10.20). For anxiety, patients that had ≥50% improvement in symptoms had significantly higher mean WAI-SR scores (mean=54.1) than those that did not (mean=47.4), *d* =0.62, t(121)=3.62, p<0.001. For depression, patients that had ≥50% improvement in symptoms had significantly higher mean WAI-SR scores (mean=53.8) than those that did not (mean=47.5), *d*=0.58, t(120)=3.36, p=0.001. Therefore, patients that had ≥50% improvement in both depressive and anxiety symptoms reported significantly higher therapeutic alliance than those that did not.

Results from logistic regression models predicting clinically significant symptoms are presented in Table 3. After controlling for demographic and baseline clinical covariates, therapeutic alliance was significantly associated with clinically significant improvement on both the GAD-7 (OR=1.08, p<0.001, 95% CI: [1.04, 1.14]) and PHQ-8 (OR=1.05, p=0.01, 95% CI: [1.01, 1.09]) scales, indicating that patients reporting higher therapeutic alliance were more likely to have clinically significant improvement than those with lower therapeutic alliance. The only other covariate reaching statistical significance was baseline symptom severity. As expected, patients with higher baseline PHQ-8 clinical symptom severity were less likely to have clinically significant improvement at follow-up than those with lower baseline severity.

**Table 3:**
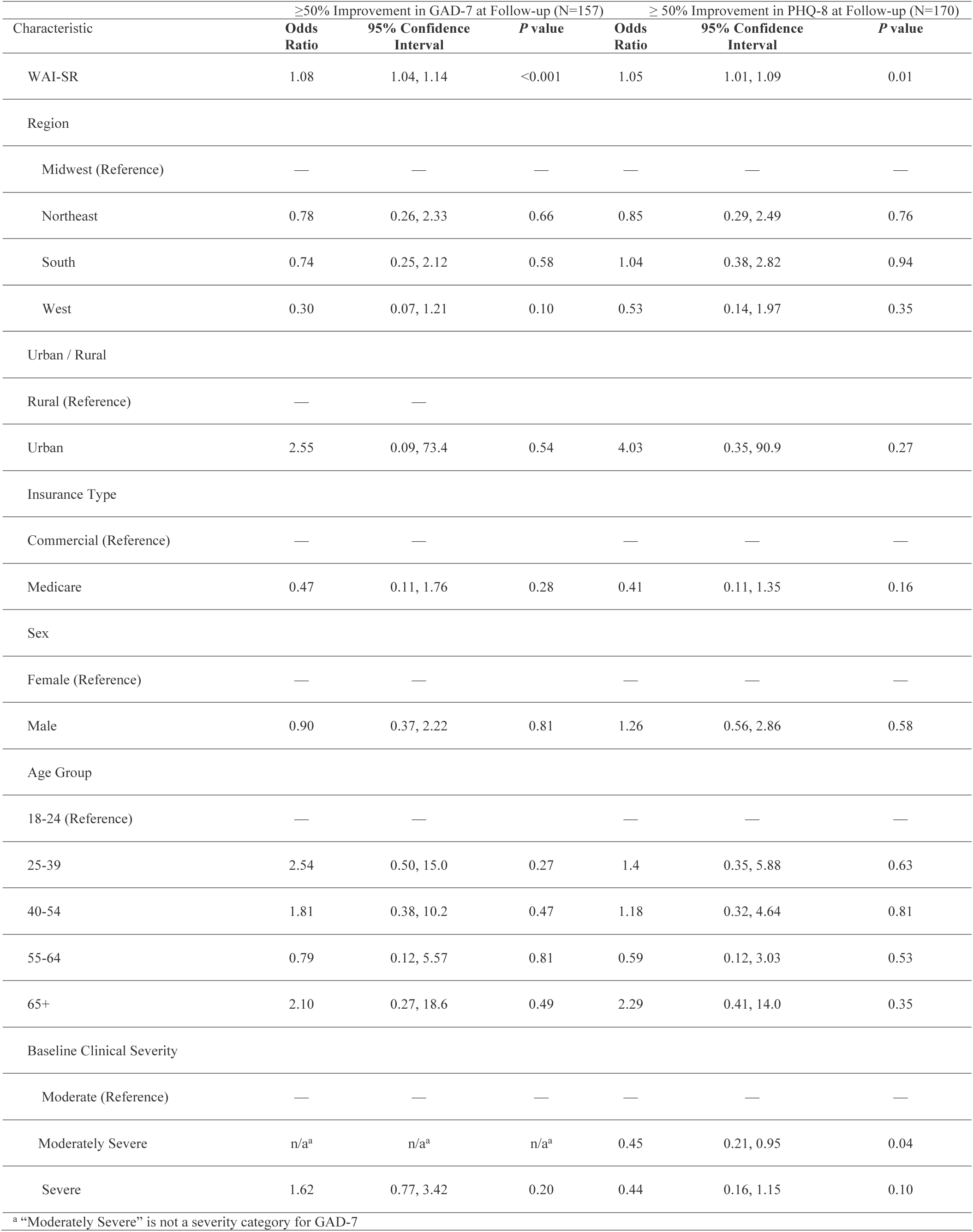
Logistic regression models predicting ≥50% improvement in symptoms at follow-up for GAD-7 and PHQ-8.

## Discussion

### Principal Results

In this retrospective study, we found clinically significant improvement among patients receiving care in a fully virtual psychiatry setting. Therapeutic alliance emerged as a significant predictor of clinical outcomes for both depression and anxiety after 8 to 16 weeks of treatment. These results have important implications for the evolving landscape of telepsychiatry care.

### Limitations

Several limitations warrant mention. Our study examined outcomes over a relatively short timeframe (8-16 weeks). Longer-term follow-up could provide additional insights into the durability of improvements. Furthermore, while therapeutic alliance emerged as the primary predictor in our model, unmeasured variables such as treatment frequency and medications may also contribute to outcomes. Future research could explore how specific components of alliance function in virtual settings and investigate potential moderators of the alliance-outcome relationship in telepsychiatry.

### Comparison With Prior Work

The robust clinical improvements observed in our virtual psychiatry cohort challenge traditional assumptions that in-person care is necessary for development of a strong therapeutic alliance and effective psychiatric treatment. Patients reported higher levels of therapeutic alliance than those measured in several recently published articles and experienced clinically meaningful symptom reduction across standardized measures^40–42^. Combined, our results add to a growing body of evidence supporting the efficacy of virtual psychiatric care^10,15,24,27^.

The identification of therapeutic alliance as the primary predictor of outcomes merits particular attention. While recent meta-analyses have found that the association between therapeutic alliance and outcomes in teletherapy is significant, there is less evidence that this finding remains true for telepsychiatry^43^. Our findings that therapeutic alliance was the only significant predictor of outcomes for both depressive and anxiety symptoms reinforce the critical importance of this relationship factor, even in telepsychiatry settings.

Taken together, our findings have several practical implications for clinical practice. First, given that therapeutic alliance has measurable and significant additive value in overall treatment, it should be emphasized as an individual contributor to successful treatment. In the present study, psychiatrists spent 20-30 minutes at each visit, using that time to develop a bond that forms the foundation of a positive therapeutic relationship. Our psychiatrists utilize an open communication style, create an environment for patient-centered care, and work on shared goals and tasks. These are time-intensive strategies that would be difficult to accomplish in shorter appointments. Second, routine assessment of therapeutic alliance may serve as a valuable early indicator of treatment trajectory, potentially allowing for targeted interventions should the alliance appear subpar. Third, psychiatric training programs should emphasize alliance building techniques tailored to virtual interactions as telepsychiatry has transitioned from an alternative solution to a permanent fixture in the psychiatric care landscape.

### Conclusions

In conclusion, this study demonstrates that fully virtual psychiatric care can produce clinically meaningful improvements, with therapeutic alliance serving as the cornerstone of effective treatment. Our findings underscore the critical importance of human connection—even when mediated through digital platforms—in achieving successful therapeutic outcomes.

## Acknowledgements

We would like to thank the psychiatrists at Talkiatry who are transforming psychiatry with accessible, human, and responsible care. We also thank Ismail Amin and Mindy Lee-Faczek for assistance with supporting alliance survey development and data collection, and Kartik Venkatachalam for comments on the manuscript. This study was sponsored by Talkiatry.

## Data Availability

The data are available from the corresponding author upon reasonable request. Only deidentified data will be made available.

## Authors’ Contributions

NP analyzed the data. CP and NP wrote this paper. CP and GG conceived this study.

---

**Figure 1:**
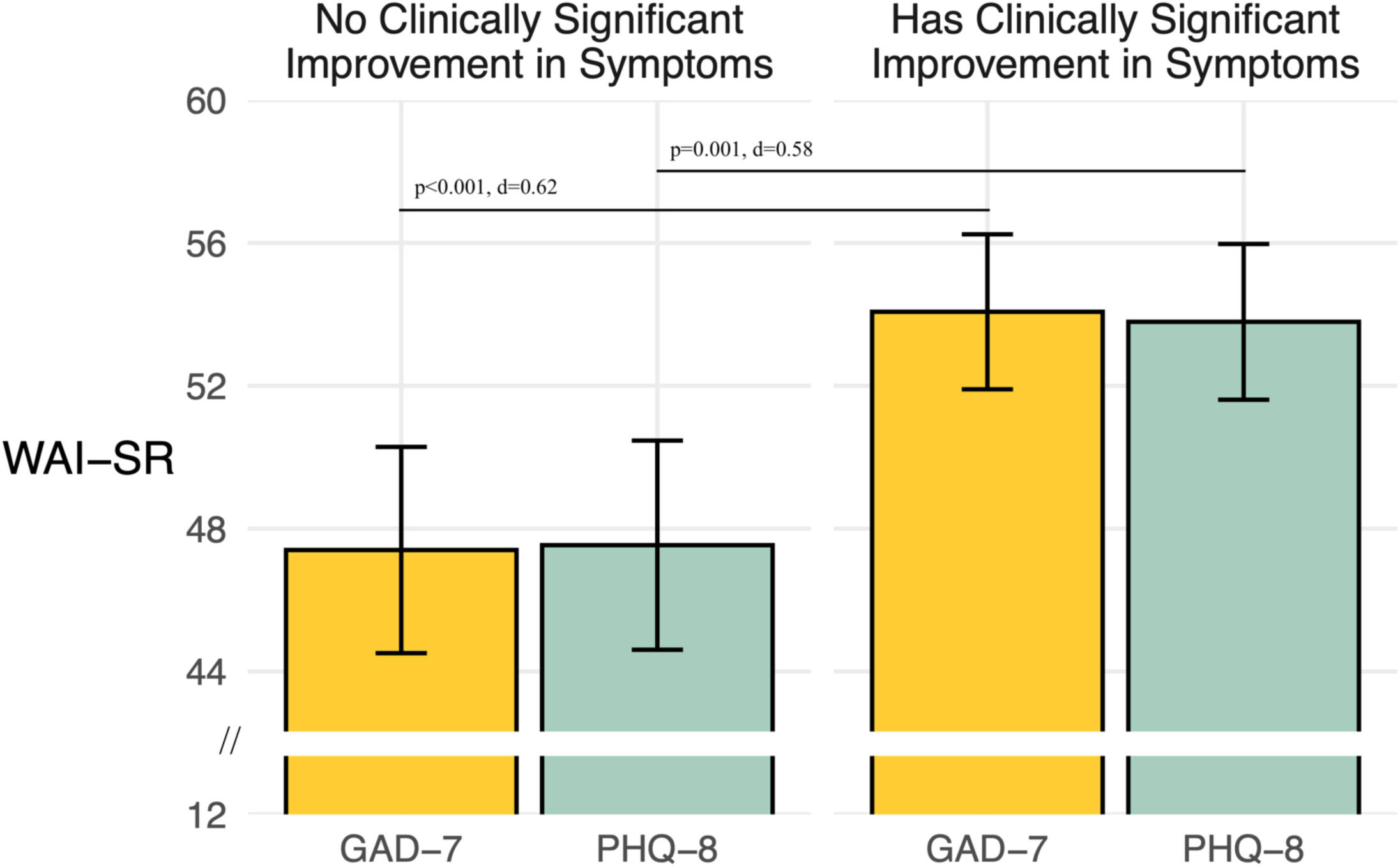
Alliance (WAI-SR) for N=201 patients separately for those who do not have clinically significant improvement in anxiety (GAD-7) or depressive (PHQ-8) symptoms and those that do at follow-up. Clinically significant improvement is defined as having ≥50% improvement in symptoms at 8-16 week follow-up relative to baseline. Higher WAI-SR values indicate stronger therapeutic alliance. Bars indicate means and error bars indicate 95% confidence intervals.

## Supplementary Materials

### WAI-SR Items

| Item | Original | Revised for Current Study |
| --- | --- | --- |
| **Prompt** | Below is a list of statements and questions about experiences people might have with their therapy or therapist. Some items refer directly to your therapist with an underlined space -- as you read the sentences, mentally insert the name of your therapist in place of _____ in the text. Think about your experience in therapy, and decide which category best describes your own experience. IMPORTANT!!! Please take your time to consider each question carefully | The following survey has a list of statements and questions about experiences people might have with their treating psychiatrist.<br>Think about your experience in treatment and decide which response best describes your own experience.<br>Please take your time answering each question and answer each as honestly as possible.<br>Your answers help us make Talkiatry better for all our patients. |
| **1** | As a result of these sessions I am clearer as to how I might be able to change. | As a result of these sessions, I am clearer as to how I might be able to change. [No change from original] |
| **2** | What I am doing in therapy gives me new ways of looking at my problem. | What I am doing in treatment gives me new ways of looking at my problem. |
| **3** | I believe _____ likes me | I believe my psychiatrist likes me. |
| **4** | _____ and I collaborate on setting goals for my therapy. | My psychiatrist and I collaborate on setting goals for my treatment. |
| **5** | _____ and I respect each other. | My psychiatrist and I respect each other. |
| **6** | _____ and I are working towards mutually agreed upon goals. | My psychiatrist and I are working towards mutually agreed upon goals. |
| **7** | I feel that _____ appreciates me. | I feel that my psychiatrist appreciates me. |
| **8** | _____ and I agree on what is important for me to work on. | My psychiatrist and I agree on what is important for me to work on. |
| **9** | I feel _____ cares about me even when I do things that he/she does not approve of. | I feel my psychiatrist cares about me even when I do things that they do not approve of. |
| **10** | I feel that the things I do in therapy will help me to accomplish the changes that I want. | I feel that the things I do in treatment will help me to accomplish the changes that I want. |
| **11** | _____ and I have established a good understanding of the kind of changes that would be good for me. | My psychiatrist and I have established a good understanding of the kind of changes that would be good for me. |
| **12** | I believe the way we are working with my problem is correct. | I believe the way we are working with my problem is correct. [No change from original] |

Goal Items: 4, 6, 8, 11; Task Items: 1, 2, 10, 12; Bond Items: 3, 5, 7, 9

## Footnotes

1 We were not able to calculate Cronbach’s alpha for PHQ-8 or GAD-7 as only the total scores, not the scores for individual items, were accessible at the time of analysis.

